# Governance, Accountability and Post-Deployment Monitoring Preferences for AI Integration in West African Clinical Practice: A Mixed-Methods Study

**DOI:** 10.64898/2026.03.30.26349782

**Authors:** Benjamin S. Chudi Uzochukwu, Yakubu Joel Cherima, Ugo Uwadiako Enebeli, Chinyere Cecelia Okeke, Adaora Chinelo Uzochukwu, Amobi Omoha, Blessing Hassan, Emmanuel Majiyebo Eronu, Shehu Mohammed Yusuf, Kennedy Anenechukwu Uzochukwu, Eziyi Iche Kalu

**Author notes:** **CORRESPONDING AUTHOR: Enebeli, Ugo Uwadiako** Department of Community Medicine, Federal University of Technology Owerri, Nigeria.

## Abstract

**Background:** The integration of artificial intelligence (AI) into clinical practice holds transformative potential for healthcare in West Africa, but safe deployment requires context-appropriate governance, accountability, and post-deployment monitoring frameworks. This cross-sectional mixed-methods study examined preferences and concerns of West African clinicians and technical experts regarding AI governance structures, post-deployment surveillance mechanisms, and accountability allocation.

**Methods:** A structured questionnaire was administered to 136 physicians affiliated with the West African College of Physicians (February 22-28, 2026), complemented by 72 key informant interviews with technical leads, AI developers, data scientists, policymakers, and healthcare leaders. Data were analyzed using descriptive statistics, inferential tests, and thematic analysis.

**Results:** Clinicians strongly preferred independent regulatory bodies (40.4%) for overseeing AI tool performance, with high trust ratings (mean:4.3/5), while vendor self-monitoring received minimal support (3.7%, mean:2.4/5). Real-time dashboards were the most favored monitoring approach (41.9%). Clear accountability pathways (94.1%), algorithm transparency (91.9%), and real-time performance data (89.7%) were rated essential by majorities. Major concerns included clinicians being unfairly blamed for AI errors (76.5%), excessive vendor control (72.8%), and absence of clear reporting pathways (69.9%). Qualitative findings emphasized continuous performance tracking for accuracy, fairness, and bias; structured incident reporting; protocols for model drift and failure; and multi-layered governance combining independent oversight, institutional AI committees, and explicit liability frameworks.

**Conclusion:** This study provides the first empirical evidence from West Africa on clinician preferences for AI governance. Findings offer actionable guidance for policymakers to build trustworthy, equitable, and safe AI integration frameworks that prioritize transparency, independent oversight, and clinician protection.

## INTRODUCTION

The use of artificial intelligence (AI) into healthcare signifies one of the most profound changes in contemporary medicine. AI is already being studied and used in complex health situations around the world, with mixed outcomes (Rech, 2025). It can predict disease outbreaks and help clinicians make decisions about diagnosis and treatment. In affluent environments, deep learning algorithms have equalled or surpassed human expert performance in tasks such as grading diabetic retinopathy and classifying skin cancer; however, the majority of evidence is retrospective and lacks substantial prospective real-world validation (Liu et al., 2019; Topol, 2019).

Global efforts to establish governance frameworks for healthcare AI are advancing rapidly. The World Health Organization (WHO) has provided leadership through its 2021 guidelines on the ethics and governance of AI for health (WHO, 2021); and the 2025 update on large multi-modal models (WHO, 2025). These documents outline six core principles: protecting autonomy, promoting human well-being, safety, sustainability, responsiveness, and accountability. A recent narrative review identified seven key governance domains: bias and fairness, explainability, safety and quality, privacy, accountability and liability, human oversight, and procurement; emphasizing that governance failures typically appear as practical problems such as miscalibrated alerts, subgroup performance cliffs, untracked model updates, opaque interfaces, and uncertainty about who can pause, roll back, or retire harmful systems (Bailo et al., 2026).

In West Africa, AI applications in healthcare are transitioning from pilot projects to real-world deployment. Notable examples include AI-enhanced cervical cancer screening by community health workers in Senegal (Rech, 2025), early cancer risk prediction models in Nigeria (Akingbola et al., 2024), and federated learning initiatives for tuberculosis chest X-ray interpretation across the region (Rech, 2025). While these demonstrate promising local innovation, the absence of established governance frameworks raises serious concerns about long-term safety, effectiveness, and equity.

The stakes are particularly high in West Africa. Health systems face heavy burdens from infectious diseases, maternal and child health challenges, and rising non-communicable diseases (Owoyemi et al., 2020). Chronic clinician shortages make AI attractive, yet poorly governed tools risk worsening the strain on overstretched workers (Rech, 2025). Variable regulatory capacity across countries heightens the danger of a “digital divide 2.0,” while linguistic, cultural, and contextual diversity increases the likelihood of poor model performance when tools are transferred across settings (Owoyemi et al., 2020; ThisDayLive, 2025).

Post-deployment monitoring is especially critical. Unlike traditional medical devices, AI models can degrade over time due to data drift or concept drift (Cuocolo et al., 2025). Without continuous surveillance, performance decline may go undetected, potentially causing harm. Trust remains central to successful AI integration. Clinicians who distrust governance structures, fear unfair blame for AI errors, or perceive excessive vendor control are less likely to adopt these tools effectively (Asan et al., 2020; Bailo et al., 2026).

Despite growing international guidance, no study has yet systematically examined West African clinicians’ preferences for AI governance, accountability mechanisms, and post-deployment monitoring. This study addresses this critical evidence gap by providing the first empirical data on how West African physicians and technical experts believe AI should be governed, monitored, and held accountable in clinical practice.

### Primary Research Question

What governance structures, accountability mechanisms, and post-deployment monitoring frameworks do West African clinicians and technical experts prefer for the safe and effective integration of artificial intelligence into clinical practice?

### Secondary Research Questions

1. Which institutional bodies (independent regulatory agencies, government ministries, hospital administrations, or vendors) do clinicians trust most to oversee AI tool performance after deployment, and what explains these preferences?
2. What specific monitoring approaches and components do clinicians and technical experts consider essential for effective post-deployment surveillance of AI systems?
3. What are the primary concerns of clinicians regarding liability, blame assignment, and reporting pathways for AI-related errors and adverse events?
4. What core components do technical experts, AI developers, and policymakers recommend for inclusion in a context-appropriate West African AI governance framework?

## METHODS

### Study design and study area

This cross-sectional mixed-methods study included a structured, self-administered questionnaire to assess the attitudes of West African professionals regarding artificial intelligence in healthcare and a qualitative exploration through key informant interviews (KIIs). The study was conducted across West African countries from February 22 to 28, 2026.

### Study Population and Sampling

The target group comprised physicians affiliated with the West African College of Physicians (WACP), the principal institution for postgraduate medical education in the sub-region. Most of the WACP fellows and members work in Nigeria; however, they can work in any West African country. This is because the college was started in and has a lot of students in Nigeria. A convenience sampling method was used with the college’s current communication channels to find willing participants. The study sought to capture a comprehensive spectrum of physician perspectives from the sub-region, imposing no restrictions on specialty, years of experience, or practice environment.

### Sample size determination

A mixed-methods approach was employed. For the quantitative phase, a cross-sectional survey was administered to physicians of the WACP. The sample size was calculated on the WinPepi sample size calculator, version 11.65 (Abramson, 2004), to estimate the primary outcome (e.g., ‘positive perception of AI utility’) with a 95% confidence level, a 5% acceptable difference, and a 10% non-response rate. Using an assumed proportion of 92% (0.92) positive perception of AI utility from a previous study (van der Meijden et al., 2023), the initial sample size was 126. This was rounded up to 136 for more precision. This sample size provides precision of approximately ±8% for proportion estimates, adequate for describing general patterns while acknowledging limitations for subgroup analyses.

For the qualitative study, a sample size of 72 was utilized. This sample size was sufficient to achieve thematic saturation across diverse stakeholder perspectives, based on established qualitative research guidelines (Braun & Clarke, 2021).

### Data Collection

A pre-test was conducted on the study tools, and the Cronbach’s alpha was utilised to determine if multi-item scales were consistent with each other and values greater than 0.70 were considered acceptable. During the survey, the questionnaire collected information on the institutional bodies (independent regulatory agencies, government ministries, hospital administrations, or vendors), clinicians trust, and what explains these preferences, what specific monitoring do clinicians and technical experts consider essential for post-deployment AI surveillance, what clinicians’ primary concerns regarding liability, blame assignment, and reporting pathways for AI-related errors are, and what technical experts and policymakers recommend including in a West African AI governance framework. The 5-point Likert scale was used to score things, with 1 being “strongly disagree” and 5 being “strongly agree”.

The Open Data Kit (ODK) platform was used to gather information, with skip patterns and validation criteria to reduce data entry errors, and the link was sent out using WACP’s email and WhatsApp communication tools. The questionnaire took 15 to 20 minutes to complete. No personally identifiable information was collected, ensuring the anonymity of participants. To gain informed consent electronically, the first page of questions had to be filled out and agreed upon before moving on.

For the qualitative study, KIIs enabled the exploration of complex phenomena, including perceptions, concerns, recommendations, monitoring mechanisms, and accountability frameworks that clinicians and technical experts prefer for the safe integration of AI into clinical practice that cannot be adequately captured through quantitative methods alone. There was a total of 72 KIIs with key stakeholders poised to offer authoritative perspectives including technical leads, AI developers, data scientists, programme directors, policymakers, ministry officials, senior clinicians, hospital leadership, and medical educators. The interviews were online and lasted less than 60 minutes using a semi-structured interview guide. For quality assurance purposes, all entries were reviewed for completeness and clarity within 24 hours.

### Data Analysis

The survey data from ODK was downloaded as an Excel file and exported into SPSS version 29, IBM Corp., Armonk, NY, USA (IBM, 2024). The descriptive statistics (frequencies, percentages, means, and standard deviations) were computed for each variable. Initially, the relationships between demographic and experiential characteristics were determined, as well as the desire to employ AI using t-tests. The ANOVA was used for inferential analysis. Factors exhibiting significant correlations (p<0.10) were included in a multivariate linear regression model to forecast the propensity to employ AI. The regression model included previous AI experience (binary), familiarity with AI (Likert scale), trust in AI information (Likert scale), perceived danger to autonomy (Likert scale), years of experience (categorical), and facility type (categorical). The cutoff for statistical significance was set at p<0.05 (two-tailed).

Data analysis for the qualitative data employed thematic analysis following the framework approach described by (Braun & Clarke, 2021). This method was selected for its flexibility and suitability for identifying, analysing, and reporting patterns within qualitative data. Analysis proceeded through several stages, namely, data familiarisation, initial coding, framework development and application of framework. The final coding framework was systematically applied to all 72 transcripts using qualitative data analysis software NVivo version 14 (Lumivero, 2026).

## RESULTS

### QUANTITATIVE RESULTS

#### Respondents’ Characteristics

A total of 137 physicians accessed the questionnaire link, resulting in 136 complete responses included in the analysis after the exclusion of one participant who declined consent. ***Table 1*** presents the demographic characteristics of the sample. The majority of respondents were from Nigeria (84.6%), with smaller representations from Ghana (5.9%), Liberia (3.6%), Togo (2.9%), Niger (1.5%), and Côte d’Ivoire (1.5%). Gender distribution was almost balanced (51.5% female, 48.5% male). The largest age group was 35-44 years (30.1%), followed by 45-54 years (26.5%). One-third of respondents (33.0%) had over 20 years of clinical experience.

**Table 1.** Demographic characteristics of survey respondents.

| Demographic Variables | Category | Frequency | Percent |
| --- | --- | --- | --- |
| Country | Nigeria | 115 | 84.6 |
|  | Ghana | 8 | 5.9 |
|  | Liberia | 5 | 3.6 |
|  | Togo | 4 | 2.9 |
|  | Niger | 2 | 1.5 |
|  | Cote d'Ivoire | 2 | 1.5 |
| Gender | Female | 70 | 51.5 |
|  | Male | 65 | 47.8 |
|  | Prefer not to say | 1 | 0.7 |
| Age group | 25-34 | 26 | 19.1 |
|  | 35-44 | 41 | 30.1 |
|  | 45-54 | 36 | 26.5 |
|  | 55-64 | 21 | 15.4 |
|  | 65+ | 11 | 8.1 |
|  | Prefer not to say | 1 | 0.7 |
| **Clinical Experience (Years)** | 0-5 | 15 | 11.0 |
|  | 6-10 | 27 | 19.9 |
|  | 11-15 | 27 | 19.9 |
|  | 16-20 | 22 | 16.2 |
|  | 20+ | 45 | 33.0 |

#### Preferred body for monitoring AI tool performance

***Table 2*** identifies who clinicians believe should be primarily responsible for overseeing AI tools after they are deployed. The largest group. This table identifies who clinicians believe should be primarily responsible for overseeing AI tools after they are deployed. The largest group, 40.4%, prefers an “Independent Body”, far exceeding support for government (20.6%) or hospital-based (32.4%) monitoring. Very few (3.7%) trust vendors to monitor themselves.

**Table 2.** Preferred body for monitoring AI tool performance.

| **Responsible body** | **Frequency** | **Percent** |
| --- | --- | --- |
| Independent body | 55 | 40.4 |
| Hospital facility | 44 | 32.4 |
| Government ministry | 28 | 20.6 |
| Vendor/manufacturing | 5 | 3.7 |
| All of the above/multiple | 2 | 1.5 |
| Other (clinicians, medical bodies | 2 | 1.5 |

#### Preferred Approach to Monitoring AI Performance

***Table 3*** shows how clinicians want monitoring information to be delivered. The most preferred method, chosen by 41.9%, is a "Real-time dashboard”, followed by monthly reports (19.9%). Annual reports received the least response (1.5%).

**Table 3.** Preferred Approach to Monitoring AI Performance.

| Monitoring Approach | Frequency (n) | Percentage (%) |
| --- | --- | --- |
| Real-time dashboard | 57 | 41.9 |
| **Weekly reports** | 12 | 8.8 |
| **Monthly reports** | 27 | 19.9 |
| **Quarterly reports** | 18 | 13.2 |
| **Annual reports** | 2 | 1.5 |
| **Multiple approaches indicated** | 20 | 14.7 |

#### Perceived Importance of Monitoring Components

***Table 4*** rates the importance of four specific monitoring components. All four are rated as "Essential/Very Important" by overwhelming majorities (ranging from 86% to 94%). Clear accountability pathways (94.1%) and transparency of algorithms (91.9%) are the top priorities.

**Table 4.** Perceived Importance of Monitoring Components.

| Monitoring Component | Essential/Very Important (%) | Moderately Important (%) | Slightly/Not Important (%) |
| --- | --- | --- | --- |
| Real-time performance data | 89.7 | 8.1 | 2.2 |
| Transparency of algorithms | 91.9 | 5.9 | 2.2 |
| Clear accountability pathways | 94.1 | 4.4 | 1.5 |
| Regular independent audits | 86.0 | 11.0 | 3.0 |

#### Clinician Trust in Different Governance Structures

***Table 5*** quantifies the level of trust clinicians placed in different potential governing bodies for AI, on a 5-point scale. An independent regulatory body received the highest trust rating (Mean=4.3), while AI vendors receive the lowest (Mean=2.4).

**Table 5.**
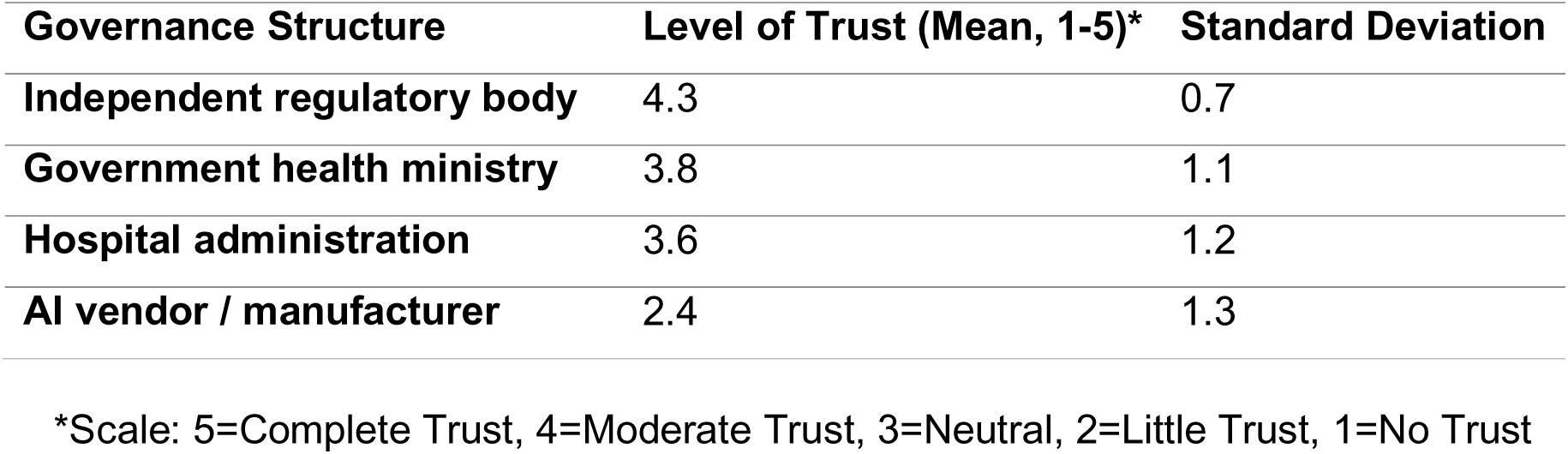
Clinician Trust in Different Governance Structures.

#### Governance and Policy Support Scale

***Table 6*** provides a statistical check for the questions related to governance and policy. The Cronbach’s Alpha of 0.79 is in the “Questionable/Acceptable” range, indicating moderate internal consistency.

**Table 6.**
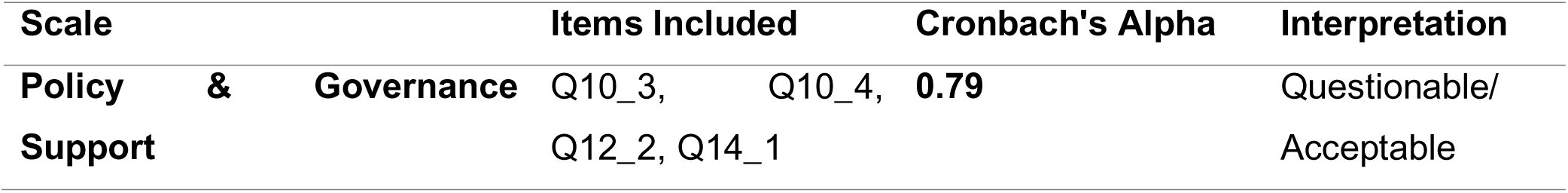
Reliability Analysis - Governance and Policy Support Scale.

| Scale | Items Included | Cronbach's Alpha | Interpretation |
| --- | --- | --- | --- |
| Policy & Governance Support | Q10_3, Q10_4, Q12_2, Q14_1 | 0.79 | Questionable/Acceptable |

#### Framework Components Preferred by Clinicians for AI governance

***Table 7*** presents clinician preferences regarding two key aspects of AI governance: the preferred body to oversee AI tools and the preferred frequency and format for receiving monitoring information. Independent oversight received the highest level of support at 40.4%. Hospital facilities were the second most preferred option at 32.4%, government ministry was preferred by 20.6% of clinicians and vendors received the lowest level of support at 3.7%,

**Table 7.** Framework Components Preferred by Clinicians for AI Governance.

| Framework component | Percent (%) |
| --- | --- |
| **Who should monitor AI?** |  |
| **Independent Oversight** | 40.4 |
| **Government Ministry** | 20.6 |
| **Hospital Facilities** | 32.4 |
| **Vendors** | 3.7 |
| **Frequency of monitoring** |  |
| **Real time dashboard** | 41.9 |
| **Weekly reports** | 8.8 |
| **Monthly reports** | 19.9 |
| **Quarterly reports** | 13.2 |
| **Annual reports** | 1.5 |

#### Essential Safety and Accountability Requirements

***Table 8*** presents clinician ratings of three framework components related to safety validation and accountability for artificial intelligence in healthcare. The data show the percentage of clinicians who rated each component as “Essential” or “Very Important” for AI governance. In terms of pre-deployment certification component, it shows that 86.0% of clinicians rate regular independent audits as “Essential” or “Very Important.” For post-deployment surveillance component, 89.7% of clinicians rate real-time performance data monitoring as “Essential” or “Very Important.” For accountability, assignment component, it shows that 94.1% of clinicians rate clear accountability pathways as “Essential” or “Very Important.” This is the highest-rated component among all framework elements.

**Table 8.** Essential Safety and Accountability Requirements.

| **Framework Component** | **Clinicians Rating as "Essential/Very Important" (%)** |
| --- | --- |
| **Pre-deployment Certification** | 86.0% |
| **Post-deployment Surveillance** | 89.7% |
| **Accountability Assignment** | 94.1% |

#### Concerns About Current Accountability Mechanisms

***Table 9*** quantifies clinician anxieties about the existing or absent systems for AI accountability. Majorities express concern that current systems cannot ensure AI accountability (68.4%) vendors have too much control (72.8%), clinicians will be unfairly blamed for AI errors (76.5%), and there is no clear pathway for reporting AI-related harm (69.9%).

**Table 9.** Concerns About Current Accountability Mechanisms.

| Concern | Agree/Strongly Agree (%) | Neutral (%) | Disagree/Strongly Disagree (%) |
| --- | --- | --- | --- |
| Current systems cannot ensure AI accountability | 68.4 | 19.1 | 12.5 |
| Vendors have too much control over monitoring | 72.8 | 16.2 | 11.0 |
| Clinicians will be unfairly blamed for AI errors | 76.5 | 13.2 | 10.3 |
| No clear pathway for reporting AI-related harm | 69.9 | 17.6 | 12.5 |

## QUALITATIVE RESULTS

A total of 72 key informant interviews were completed as in ***Table 10*** from all stakeholder groups including Technical Leads, AI Developers, Data Scientists, and Policymakers.

**Table 10.** Categories of key informants.

| Informant Category | Number of Participants |
| --- | --- |
| Technical Leads | 15 |
| AI Developers | 9 |
| Data Scientists | 16 |
| Program Directors | 7 |
| Policymakers | 6 |
| Ministry Officials | 6 |
| Senior Clinicians | 3 |
| Hospital Leadership | 4 |
| Department Heads | 2 |
| Medical Educators | 4 |

### The Indispensable Need for Continuous Monitoring

Technical experts were unanimous that: *“Deployment is not the finish line. It’s the starting line.*” And that a framework must be in place from day one to answer four key questions: Is it still accurate? Is it still fair? Is it still safe? Is it still improving care?

### Core Components of a post-deployment monitoring framework

Respondents provided a detailed and structured approach to monitoring; for example, that continuous performance tracking was non-negotiable. According to one of the respondents, it involved regularly tracking *“accuracy, sensitivity, specificity, false positives/negatives, and calibration”* against real-world outcomes. They noted that monitoring needs to be stratified to detect *“emerging bias.”* Performance must be tracked across *age, gender, geographic location (urban vs. rural), and facility type… to ensure the AI isn’t performing poorly”* for specific populations. One respondent noted that *“aggregated metrics hide localized harm.”* They noted that systems must be in place to automatically detect *“data drift”* (changes in input data, e.g., a new disease outbreak), *“concept drift”* (changes in the relationship between data and outcome, e.g., a new treatment guideline), and that this required *“automated alerts”* that trigger investigation when performance drops below a threshold.

Respondents reported that user feedback and incident reporting were key, monitoring needs to be human-centred, structured *“clinician feedback loops”* to report errors or usability issues, and a formal *“incident-reporting mechanism”* to log near-misses and adverse events were essential. Additionally, that post-deployment monitoring framework must include a schedule for *“periodic re-validation using updated local data;”* and when drift is detected, the model must be *“retrained, recalibrated, or, in severe cases, retired.”*

### Governance, Accountability, and Regulatory Frameworks

Strong governance was seen as the bedrock of safe use of AI tools in healthcare. According to several respondents, key requirements include “*clinical validation”* (requiring rigorous testing and validation of AI systems before clinical use); “*data protection”* (strict compliance with national data protection laws, e.g., Nigeria’s NDPA), including consent, anonymization, and security); “*accountability and liability”* (clearly established decision-making processes and accountability in AI healthcare-related errors); *transparency* (mandating “transparency in AI decision-making processes and data use,” with developers disclosing how systems are trained and validated), *institutional governance structures* (hospitals need new structures to oversee AI. This includes an “AI governance committee” comprising hospital leadership, IT specialists, senior clinicians, legal/compliance officers, and data protection officers), and *policy priorities for governments.* Policymakers outlined a clear set of priorities including patient safety and regulatory oversight by establishing strong approval and monitoring systems, data governance and protection by enforcing robust data protection laws and cybersecurity standards and equitable access and capacity building through investment in public infrastructure and training the workforce to ensure AI benefits all.

### Handling Failure: Protocols for When AI Underperforms

Respondents noted that a mature governance framework includes clear protocols for failure. The response must be swift and structured and should include:

1. *Suspend or Restrict Use:* The priority is to prevent patient harm. The tool should be immediately paused or restricted.
2. *Conduct Root-Cause Analysis:* Investigate whether the issue is due to data pipeline problems, workflow mismatch, model drift, or population shift.
3. *Apply Corrective Actions:* Based on the analysis, “recalibrate, retrain, or redesign” the model.
4. *Communicate Transparently:* Inform clinicians and relevant authorities about the issue and the corrective steps taken. This transparency is crucial for “protecting clinician trust.”

### The key governance recommendations by the respondents include

1. Establish an independent, multistakeholder regulatory body
2. Mandate real-time performance dashboards for all deployed AI
3. Create clear legal liability frameworks
4. Develop standardized reporting protocols for AI-related incidents
5. Require regular independent audits

## DISCUSSION

This mixed-methods study investigated West African clinicians’ preferences for AI governance, accountability mechanisms, and post-deployment monitoring frameworks. The quantitative survey of 136 physicians, predominantly from Nigeria, was complemented by 72 KIIs with technical experts, policymakers, and healthcare leaders. The findings revealed four major themes: a strong preference for independent oversight over government, hospital, or vendor control; real-time dashboards as the preferred monitoring mechanism; accountability concerns dominating clinician anxieties; and recommendations for a multi-layered governance framework combining independent bodies, institutional committees, and clear liability rules.

### Governance Preferences

This study provides novel empirical evidence on AI governance preferences in West African contexts, where regulatory capacity is often limited and tools developed elsewhere may be deployed without local validation (Finkelstein et al., 2024). Prior research has focused mainly on high-income countries with mature systems (Reddy et al., 2020), while studies from low- and middle-income settings have largely been limited to commentary and position papers (Cresswell et al., 2020).

Clinicians demonstrated awareness of regional vulnerabilities and demanded independent oversight, local validation, and clear accountability pathways. These preferences align with WHO guidance, which stresses appropriate regulation for safety, accuracy, and efficacy as universal principles (WHO, 2021). Policymakers’ emphasis on equitable access and capacity building echoes the concept of “health data poverty” (Ibrahim et al., 2021) and the risk of a “digital divide 2.0” (ThisDayLive, 2025).

Quantitatively, 40.4% of clinicians preferred an independent body for overseeing AI tool performance, the most selected option, with independent regulatory bodies receiving the highest trust rating (mean 4.3/5) and vendors the lowest (mean 2.4/5). This reflects a desire for impartial oversight free from commercial interests or institutional politics. The finding is consistent with high-income country studies: 62% of UK clinicians favoured independent regulation (Castagno & Khalifa, 2020), and US physicians expressed concerns about commercial influence (Habli et al., 2020). The lower preference rate here (40.4% vs 62%) likely stems from the study offering four response options rather than a binary choice.

Low trust in vendors indicates strong scepticism of commercial self-regulation, aligning with qualitative concerns about excessive vendor control and the need for clear decision-making processes in AI-related errors (WHO, 2021). Moderate trust in government highlights both opportunities and the need for capacity building, potentially reflecting contextual factors such as regulatory capture or limited enforcement in West Africa (Akingbola et al., 2024; Owoyemi et al., 2020). This extends the literature by showing how local institutional trust dynamics shape governance preferences in resource-constrained settings (Scott et al., 2021).

### Monitoring Preferences

Clinicians strongly preferred real-time dashboards (41.9%), indicating a desire to be active participants in safety surveillance rather than passive recipients of periodic reports. Technical experts reinforced this by stressing that monitoring must continuously address four questions: “Is it still accurate? Is it still fair? Is it still safe? Is it still improving care?” This aligns with the FDA’s “total product lifecycle” approach to AI oversight (U.S. FDA, 2021). Emphasis on algorithm transparency and explainability supports existing evidence that these factors are key determinants of clinician trust (Ahmad et al., 2021; Asan et al., 2020). The study extends prior work by specifying not only what information is needed but how and when it should be delivered, with important implications for monitoring infrastructure in West Africa.

Qualitative findings highlighted the need for stratified monitoring to detect emerging bias across age, gender, location (urban vs rural), and facility type. Experts noted that “aggregated metrics hide localized harm,” a concern well-documented in the literature (Civaner et al., 2022; Ibrahim et al., 2021; Obermeyer et al., 2019), and consistent with FDA guidance on differential performance across subgroups (U.S. FDA, 2021).

### Accountability Concerns

Accountability was the dominant concern, rated essential or very important by 94.1% of clinicians: the highest priority. Major anxieties included fear of being unfairly blamed for AI errors (76.5%), excessive vendor control (72.8%), and absence of clear reporting pathways (69.9%). These findings address the “responsibility gap” in the literature, whereby liability often defaults to clinicians under existing malpractice doctrines when no one is fully responsible for AI outcomes (Price et al., 2019; van der Meijden et al., 2023). Without clear statutory allocation, clinicians may face disproportionate risk, discouraging appropriate AI use (Mello & Wang, 2020).

Qualitative participants similarly called for clearly established accountability in AI-related errors. The near-universal demand for accountability (94.1%) highlights that this is a core requirement for clinician acceptance of AI. The lack of clear reporting mechanisms is a critical regulatory gap as errors go undocumented and learning opportunities are lost (Ocloo et al., 2021; Sujan et al., 2019). In West African settings with less developed medico-legal frameworks, addressing this gap before widespread deployment is particularly urgent (Owoyemi et al., 2020).

### Implications for Policymakers

The findings provide clear direction for policymakers tasked with developing AI governance frameworks in West Africa:

#### Establish independent, multi-stakeholder regulatory bodies

The preference for independent oversight and high trust in independent bodies indicate that credibility requires perceived objectivity. Policymakers should establish regulatory authorities with representation from clinicians, data scientists, patient advocates, and other stakeholders, ensuring independence from both commercial interests and political influence (WHO, 2021).

#### Mandate real-time performance dashboards for deployed AI

The strong preference for real-time monitoring dashboards indicates that clinicians want immediate access to performance data. Policymakers should require that deployed AI systems provide accessible, real-time dashboards enabling clinicians to track accuracy, fairness, and safety continuously as noted elsewhere (Cuocolo et al., 2025).

#### Create clear liability frameworks protecting clinicians

The near-universal demand for accountability pathways in this study and widespread fear of unfair blame indicate that liability frameworks must be established before AI deployment. Legislation should clearly allocate responsibility among developers, deployers, and users, protecting clinicians from bearing liability for errors from algorithmic failures (Price et al., 2019).

#### Develop standardized reporting protocols for AI-related incidents

The finding that a majority see no clear pathway for reporting AI-related harm indicates an urgent need for incident reporting systems. Policymakers should establish standardized protocols enabling clinicians to report near-misses and adverse events, treating these as learning opportunities for system improvement (Sujan et al., 2019).

### Limitations

This study has several limitations that should be considered when interpreting the findings. First, the geographic representation was heavily skewed toward Nigeria (84.6% of survey respondents), limiting generalizability to other West African countries with different health systems, regulatory environments, and cultural contexts. Future research should include more balanced representation across Francophone and Lusophone West African nations, second, the convenience sampling method through WACP communication channels may have introduced selection bias, third, the study measured preferences and attitudes rather than actual behaviour.

## CONCLUSION

This study provides the first empirical evidence on West African clinician preferences for AI governance, accountability mechanisms, and post-deployment monitoring. The findings reveal a clear mandate for independent oversight, real-time transparency, and explicit liability frameworks. Clinicians distrust vendor self-regulation and fear being unfairly blamed for AI errors. Technical experts emphasize continuous monitoring, bias surveillance, structured failure protocols, and multi-layered governance combining independent bodies, institutional committees, and clear accountability pathways.

The convergence of quantitative and qualitative evidence strengthens confidence in these findings and provides actionable guidance for policymakers. An independent, multi-stakeholder regulatory body, mandated real-time performance dashboards, clear liability frameworks, standardized incident reporting protocols, and institutional AI governance committees represent the core components of a governance framework aligned with clinician trust and safety priorities. Without such frameworks, the promise of AI in West African healthcare risks being undermined by clinician scepticism, regulatory gaps, and avoidable patient harm. The preferences and concerns documented here provide a roadmap for building governance structures that can earn trust, ensure safety, and enable the equitable integration of AI into clinical practice across the region. The evidence from this study demonstrates that West African clinicians are ready to engage with AI, but only under governance frameworks that prioritize accountability, transparency, and patient safety above all else.

## DECLARATIONS

### Ethical approval

Ethical approval was obtained from the Ethical Clearance Committee of the University of Nigeria Teaching Hospital (ethical clearance number UNTH/HREC/2026/01/4157). The study was conducted in accordance with the Declaration of Helsinki. Electronic informed consent was obtained from all participants. For the survey, consent was implied by voluntary completion of the questionnaire after reading the participant information sheet on the first page of the ODK form. For key informant interviews, written informed consent was obtained electronically prior to the interview. No personally identifiable information was collected.

### Data availability

The de-identified quantitative survey dataset and de-identified qualitative transcripts are available from the corresponding author upon reasonable request and subject to institutional data-sharing approval.

### Funding

This research received no specific grant from any funding agency in the public, commercial, or not-for-profit sectors.

### Authors’ contributions

BSCU: Conceptualization, Methodology, Supervision, Writing - review & editing. YJC: Data curation, Formal analysis, Writing - original draft. UUE: Conceptualization, Methodology, Investigation, Project administration, Writing - original draft, Writing - review & editing (corresponding author). CCO: Investigation, Resources, Writing - review & editing. ACU: Validation, Writing - review & editing. AO: Investigation, Writing - review & editing. BH: Data curation, Writing - review & editing. EME: Software, Visualization, Writing - review & editing. SMY: Software, Formal analysis, Writing - review & editing. KAU: Investigation, Writing - review & editing. EIK: Validation, Writing - review & editing. All authors read and approved the final manuscript and agree to be accountable for all aspects of the work.

### Conflicts of interest / Competing interests

The authors declare no competing interests.

## Notes

### Competing Interest Statement

The authors have declared no competing interest.

### Author Declarations

Ethical approval was obtained from the Ethical Clearance Committee of the University of Nigeria Teaching Hospital (ethical clearance number UNTH/HREC/2026/01/4157).

## REFERENCES

Abramson, J. H. (2004). WINPEPI (PEPI-for-Windows): Computer Programs for Epidemiologists. Epidemiologic Perspectives & Innovations, 1(1), 6. 10.1186/1742-5573-1-6

Ahmad, Z., Rahim, S., Zubair, M., & Abdul-Ghafar, J. (2021). Artificial intelligence (AI) in medicine, current applications and future role with special emphasis on its potential and promise in pathology: present and future impact, obstacles including costs and acceptance among pathologists, practical and philosop…. Diagnostic Pathology, 16(1), 24-. 10.1186/s13000-021-01085-4

Akingbola, A., Adegbesan, A., Ojo, O., Otumara, J. U., & Alao, U. H. (2024). Artificial Intelligence And Cancer Care in Africa. *Journal of Medicine*, Surgery, and Public Health, 3(3), 100132. 10.1016/j.glmedi.2024.100132

Asan, O., Bayrak, A. E., & Choudhury, A. (2020). Artificial Intelligence and Human Trust in Healthcare: Focus on Clinicians. Journal of Medical Internet Research, 22(6), e15154. 10.2196/15154

Bailo, P., Nittari, G., Pesel, G., Basello, E., Spasari, T., & Ricci, G. (2026). Governing Healthcare AI in the Real World: How Fairness, Transparency, and Human Oversight Can Coexist: A Narrative Review. Sci, 8(2), 36. 10.3390/sci8020036

Braun, V., & Clarke, V. (2021). One size fits all? What counts as quality practice in (reflexive) thematic analysis? Qualitative Research in Psychology, 18(3), 328–352. 10.1080/14780887.2020.1769238

Castagno, S., & Khalifa, M. (2020). Perceptions of Artificial Intelligence Among Healthcare Staff: A Qualitative Survey Study. Frontiers in Artificial Intelligence, 3, 578983. 10.3389/frai.2020.578983

Civaner, M. M., Uncu, Y., Bulut, F., Chalil, E. G., & Tatli, A. (2022). Artificial intelligence in medical education: a cross-sectional needs assessment. BMC Medical Education, 22(1), 772. 10.1186/s12909-022-03852-3

Cresswell, K., Callaghan, M., Khan, S., Sheikh, Z., Mozaffar, H., & Sheikh, A. (2020). Investigating the use of data-driven artificial intelligence in computerised decision support systems for health and social care: A systematic review. Health Informatics Journal, 26(3), 2138–2147. 10.1177/1460458219900452

Cuocolo, R., Bernardini, D., Pinto dos Santos, D., Klontzas, M. E., Akinci D’Antonoli, T., Semedo, L. C., Decoster, R., Huisman, M., Kotter, E., Martí-Bonmatí, L., Minoiu, C., Neri, E., Nikolaou, K., Radzina, M., Sala, E., Shelmerdine, S. C., Topff, L., & Williams, M. C. (2025). AI medical device post-market surveillance regulations: consensus recommendations by the European Society of Radiology. Insights into Imaging, 16(1), 275-. 10.1186/s13244-025-02146-8

Finkelstein, J., Gabriel, A., Schmer, S., Truong, T. T., & Dunn, A. (2024). Identifying Facilitators and Barriers to Implementation of AI-Assisted Clinical Decision Support in an Electronic Health Record System. Journal of Medical Systems, 48(1), 89. 10.1007/s10916-024-02104-9

Habli, I., Lawton, T., & Porter, Z. (2020). Artificial intelligence in health care: accountability and safety. Bulletin of the World Health Organization, 98(4), 256. 10.2471/BLT.19.237487

IBM. (2024). *IBM SPSS Statistics 29*. IBM Support. https://www.ibm.com/support/pages/downloading-ibm-spss-statistics-29

Ibrahim, H., Liu, X., Zariffa, N., Morris, A. D., & Denniston, A. K. (2021). Health data poverty: an assailable barrier to equitable digital health care. The Lancet Digital Health, 3(4), e260–e265. 10.1016/S2589-7500(20)30317-4

Liu, X., Faes, L., Kale, A. U., Wagner, S. K., Fu, D. J., Bruynseels, A., Mahendiran, T., Moraes, G., Shamdas, M., Kern, C., Ledsam, J. R., Schmid, M. K., Balaskas, K., Topol, E. J., Bachmann, L. M., Keane, P. A., & Denniston, A. K. (2019). A comparison of deep learning performance against health-care professionals in detecting diseases from medical imaging: a systematic review and meta-analysis. The Lancet Digital Health, 1(6), e271–e297. 10.1016/S2589-7500(19)30123-2

Lumivero. (2026, January 27). *About NVivo (NVivo 14 Windows)*. Lumivero Community. https://community.lumivero.com/s/article/TRC-About-NVivo-NVivo-14-Windows?language=en_US

Mello, M. M., & Wang, C. J. (2020). Ethics and governance for digital disease surveillance. Science (New York, N.Y.), 368(6494), 951–954. 10.1126/science.abb9045

Obermeyer, Z., Powers, B., Vogeli, C., & Mullainathan, S. (2019). Dissecting racial bias in an algorithm used to manage the health of populations. *Science (New York*, N.Y.), 366(6464), 447–453. 10.1126/science.aax2342

Ocloo, J., Garfield, S., Franklin, B. D., & Dawson, S. (2021). Exploring the theory, barriers and enablers for patient and public involvement across health, social care and patient safety: a systematic review of reviews. Health Research Policy and Systems, 19(1), 8. 10.1186/s12961-020-00644-3

Owoyemi, A., Owoyemi, J., Osiyemi, A., & Boyd, A. (2020). Artificial Intelligence for Healthcare in Africa. Frontiers in Digital Health, 2, 6. 10.3389/fdgth.2020.00006

Price, W. N., Gerke, S., & Cohen, I. G. (2019). Potential Liability for Physicians Using Artificial Intelligence. JAMA, 322(18), 1765–1766. 10.1001/jama.2019.15064

Rech, D. (2025, September 15). The Ethics of AI-Driven Health Projects in Africa. Think Global Health. https://www.thinkglobalhealth.org/article/the-ethics-of-ai-driven-health-projects-in-africa

Reddy, S., Allan, S., Coghlan, S., & Cooper, P. (2020). A governance model for the application of AI in health care. Journal of the American Medical Informatics Association, 27(3), 491–497. 10.1093/jamia/ocz192

Scott, I. A., Carter, S. M., & Coiera, E. (2021). Exploring stakeholder attitudes towards AI in clinical practice. BMJ Health & Care Informatics, 28(1), e100450. 10.1136/bmjhci-2021-100450

Sujan, M., Furniss, D., Grundy, K., Grundy, H., Nelson, D., Elliott, M., White, S., Habli, I., & Reynolds, N. (2019). Human factors challenges for the safe use of artificial intelligence in patient care. BMJ Health & Care Informatics, 26(1), e100081. 10.1136/bmjhci-2019-100081

ThisDayLive. (2025, September 25). *Parliamentarians Urged to Champion AI Adoption in West African Healthcare*. This Day Newspaper. https://www.thisdaylive.com/2025/09/25/parliamentarians-urged-to-champion-ai-adoption-in-west-african-healthcare/

Topol, E. J. (2019). High-performance medicine: the convergence of human and artificial intelligence. Nature Medicine, 25(1), 44–56. 10.1038/s41591-018-0300-7

U.S. FDA. (2021). *Artificial Intelligence/Machine Learning (AI/ML)-Based Software as a Medical Device (SaMD) Action Plan*. U.S. Food & Drug Administration. www.fda.gov

van der Meijden, S. L., de Hond, A. A. H., Thoral, P. J., Steyerberg, E. W., Kant, I. M. J., Cinà, G., & Arbous, M. S. (2023). Intensive Care Unit Physicians’ Perspectives on Artificial Intelligence-Based Clinical Decision Support Tools: Preimplementation Survey Study. JMIR Human Factors, 10(1), e39114. 10.2196/39114

WHO. (2021). Ethics and governance of artificial intelligence for health: WHO guidance. In Https://Iris.Who.Int/Bitstream/Handle/10665/350567/9789240037403-Eng.Pdf. World Health Organization. https://iris.who.int/handle/10665/341996

WHO. (2025). *Ethics and governance of artificial intelligence for health: Guidance on large multi-modal models*. World Health Organization. https://www.who.int/publications/i/item/9789240084759

